# Cortical activity during preparation and execution of balance recovery behavior in people after mild traumatic brain injury: A preliminary investigation

**DOI:** 10.64898/2026.03.30.26349748

**Authors:** Jacqueline A. Palmer, Keith Lohse, Peter C. Fino

**Affiliations:** Division of Physical Therapy and Rehabilitation Science, Medical School, University of Minnesota, Minneapolis, MN, United States of America; Program in Physical Therapy, Washington University School of Medicine in Saint Louis; Department of Neurology, Washington University School of Medicine in Saint Louis; Department of Health & Kinesiology, University of Utah, Salt Lake City, UT, United States of America; Department of Physical Therapy & Athletic Training, University of Utah, Salt Lake City, UT, United States of America; Department of Mechanical Engineering, University of Utah, Salt Lake City, UT, United States of America; Department of Biomedical Engineering, University of Utah, Salt Lake City, UT, United States of America

**Author notes:** Correspondence: Jacqueline A Palmer, 420 Delaware St. SE (MMC 388), Minneaopolis, MN 55455.

## Abstract

**Background and purpose:** People after mild traumatic brain injury (mTBI) show persistent deficits in reactive balance. Cortical processes engaged during preparation and execution of balance reactions are reflected in distinct cortical activity signatures that can be measured with electroencephalography (EEG). The purpose of this study was to 1) compare preparatory cortical beta activity and evoked cortical N1 responses during balance recovery in people with mTBI and controls, and 2) explore relationships between preparatory and evoked cortical activity.

**Methods:** Participants (age 21-35 years) with symptomatic mTBI (n=5, 27±13 days post-injury) and controls (n=5) completed the instrumented and modified push & release tests of reactive balance. Cortical activity was recorded using encephalography (EEG). Main outcome measures were 1) preparatory sensorimotor cortical beta-bust power and duration prior to balance perturbation onset (-1s – 0s), and 2) cortical N1 response amplitude and latency during the post-perturbation balance recovery (50-250ms).

**Results:** People with mTBI exhibited lower preparatory beta-burst power compared to controls (p=0.044, g=1.18). During balance recovery, cortical N1 responses occurred earlier in people with mTBI compared to controls (p=0.045, g=3.28). Relationships between preparatory and evoked cortical activity were altered after mTBI compared to controls; people after mTBI with greater beta-burst power and longer duration elicited shorter N1 latencies (r’s>−0.77, p’s<0.010).

**Discussion and conclusion:** The results serve as preliminary, hypothesis-generating observations to guide future research directions investigating neural signatures of reactive balance deficits in people after mTBI. The preparatory brain state before reactive balance recovery should be explored as a potential target for post-mTBI balance rehabilitation.

## 1. Introduction

Mild traumatic brain injury (mTBI) commonly causes persistent balance dysfunction, yet no reliable clinical or neuroimaging markers exist to diagnose the neural basis of these deficits or predict recovery. Between 20% and 60% of individuals report symptoms persisting beyond three months post-injury, and up to 25% report dizziness or imbalance one year after their mTBI.^1,2^ These balance impairments can affect everyday activities, increase fall risk, and reduce physical activity and independence.^3^ mTBI is a non-focal injury with diffuse effects across multiple brain regions, presenting significant challenges for characterization with structural MRI-based approaches.^4–6^ As a result, treatment is guided almost entirely by patient-reported symptoms. The most recent Clinical Practice Guidelines for physical therapy evaluation and treatment after concussion found insufficient evidence to support a clear set of motor function measures for individuals who have experienced a concussive event.^3^ This lack of objective markers is particularly problematic in rehabilitation settings, which are often the primary line of intervention for patients affected by mTBI. Functional neuroimaging approaches that can capture neural activity during functional balance behaviors relevant to clinical symptoms may hold promise to inform rehabilitation strategies.

Reactive balance, the ability to recover stability after an external perturbation, is impaired after mTBI,^7–9^ and deficits may originate from either an impaired ability to prepare motor responses or an impaired ability to execute appropriate responses. Before a loss of balance, motor plans are prepared by preselecting a reduced set of responses based on the environment and prior experience; after a loss of balance, a motor plan is rapidly selected and enacted based on available sensory information.^10,11^ Recent work indicates that people with chronic, symptomatic mTBI take longer to regain stability after a perturbation but do not take longer to initiate a reactive step compared to controls,^7^ suggesting that impaired reactive balance after mTBI is not simply due to slowed response times but may instead reflect imprecise or poorly calibrated stepping responses. Emerging mobile neuroimaging studies using functional near-infrared spectroscopy (fNIRS) have demonstrated increased prefrontal cortex activity during gait in people with persistent symptoms after mTBI,^12^ providing initial evidence that cortical processes are altered during balance and mobility tasks. However, hemodynamic-based neuroimaging cannot distinguish between preparatory and execution phases of balance control that occur on millisecond timescales. Neuroimaging modalities such as electroencephalography (EEG) offer the temporal resolution necessary to examine distinct neural processes occurring before and after balance perturbations.

Cortical beta oscillations and the balance perturbation-evoked N1 response are two EEG-derived neural signatures that reflect clinically relevant processes of sensory gating and error detection during balance control. Pre-stimulus beta oscillations (13-30 Hz) suppress perception of somatosensory stimuli, a process termed “sensory gating” that may prevent the central nervous system from being overwhelmed by predictable afferent feedback.^13^ During balance perturbations, higher pre-perturbation beta power is associated with reduced perceptual awareness of perturbation direction without compromising motor performance, suggesting effective filtering of task-irrelevant sensory information.^14^ Sensory hypersensitivity is a symptom after mTBI,^15^ and impaired sensory gating could contribute to both sensory symptoms and balance dysfunction. Following a balance perturbation, a large negative potential termed the cortical N1 response is evoked over central midline regions approximately 50-250 ms post-perturbation,^16^ localized to the supplementary motor area.^17,18^ The N1 is thought to reflect sudden detection of an error between predicted and actual sensorimotor states and the initiation of rapid balance error processing.^19,20^ Examining both preparatory beta activity and evoked N1 responses may reveal how cortical processes of sensory gating and error detection interact during reactive balance after mTBI.

The purpose of this preliminary study was to examine cortical activity during each the preparation and execution phases of reactive balance recovery in people after mTBI compared to age-matched controls. We characterized preparatory sensorimotor beta-burst activity prior to perturbation onset and the evoked cortical N1 response during balance recovery, and explored relationships between these preparatory and execution-related neural signatures. Participants completed an instrumented Push and Release paradigm, a modified version of the clinical test Push and Release (i.e., compensatory stepping from the mini Balance Evaluation Systems Test; mini-BESTest) test,^21–23^ enabling examination of cortical activity during a task that approximates clinical balance assessment. Given the small sample size, these findings are intended as preliminary, hypothesis-generating observations to inform future research on cortical mechanisms underlying reactive balance dysfunction after mTBI.

## 2. Materials and Methods

### 2.1. Participants

Ten participants (n=5 mTBI, n=5 controls) participated in this study (**Table 1**). Inclusion criteria were (1) aged 18-50 years, (2) able to walk unassisted, (3) willing and able to provide informed consent. Additional inclusion criteria for people with mTBI were (1) a clinical diagnosis of mTBI or concussion by a physician between 14-60 days prior to enrollment and (2) still reporting symptoms related to that mTBI at the time of testing. Exclusion criteria for all participants were (1) central or peripheral nervous system dysfunction (e.g.,Parkinson’s disease, vestibular schwannoma, etc.), (2) recent (<6 months) orthopedic injury that may impact balance and gait, (3) pregnancy in females (balance considerations), (4) inability or unwillingness to comply with instructions, (5) weight over 300lbs (safety considerations), (6) major reconstructive surgery to the ankle, knee, or hip, (7) diagnosed learning or mental disability, or (8) skin allergy that would react to the conductive gel. Additionally, healthy control participants were excluded if they (1) experienced an mTBI within the past year or 2) self-reported symptoms consistent with an mTBI or imbalance. Participants were identical to those reported in a previous study examining reactive balance during walking.^24^ The University of Utah Institutional Review Board approved this protocol (IRB#: 00117220) and all participants provided written informed consent. Our sample size, N=10, was determined based on resource constraints from our preliminary funding and the relative cost of collecting these EEG measures in a clinical population. We emphasize that the present study had no specific *a priori* hypotheses it was designed to strictly test, and we would lack statistical power to do so in the current sample even if we did. We emphasize that the goal of this exploratory work is hypothesis generation – providing information about ranges of effects sizes and uncertainty in the data – that can be useful in the design of future (hypothesis testing) experiments.

**Table 1.** Descriptive statistics of each group. Values are reported as mean (SD) unless otherwise noted.

|  | mTBI | controls |
| --- | --- | --- |
| Age (yrs)<sup>∞</sup> | 24.0 (5.1) | 21.8 (1.0) |
| Sex (F/M)<sup>∞</sup> | 2F/3M | 3F/1M |
| Height (cm) | 166.9 (7.4) | 169.3 (3.4) |
| Mass (kg) | 61.0 (8.3) | 64.8 (6.6) |
| Post-Concussion Symptom Scale<br>Total Symptom Severity<sup>#</sup> | 17 (7-24) | 2.5 (0-11) |
| Number of previous mTBIs<sup>#</sup> | 2 (1-5) | 0 (0-3) |
| Time since most recent mTBI<br>(days)<sup>#</sup> | 22 (18-47) | 3029* |
| Reactive step latency (ms) | 249 (103) | 176 (36) |
| Time to Stability (ms) | 1041 (211) | 975 (243) |

<sup>#</sup>reported as median (range). <sup>∞</sup>self-reported; \*only one control reported previous concussions

### 1.1. Clinical assessments

All participants completed Post-Concussion Symptom Scale (PCSS) and a structured interview to identify previous history of mTBI using the Ohio State TBI Identification Method.

### 1.2. Reactive balance paradigm

Each participant completed an instrumented, modified version of the clinical Push and Release test^22^ (i.e., compensatory stepping from the mini-BESTest). Briefly, participants were instructed to lean into an administrator’s hands until they were off-balance. Participants leaned in four directions (forward, backward, left, right) a plank-like, straight body position for each direction into an administrator’s hands until the administrator identified an inflection point where the participant’s center of mass was outside their base of support. After holding the participant in this position for 3-5 seconds, the administrator released the participant, allowing them to begin falling and to regain their balance on their own. The participant was instructed to regain their balance and avoid a fall by whatever means necessary, including taking a step or steps, after release. This procedure was repeated in all four directions (forward, backward, left, and right) in two conditions: with eyes open and with eyes closed. During the eyes closed condition, participants were instructed to close their eyes after the administrator leaned the participants, but before the release. Participants were allowed to open their eyes upon the release of support. Participant’s stance width was standardized for using a foot plate that was 8 inches (20.32 cm) long, 5.75 inches (14.61 cm) wide on the toe side, and 4 inches (10.16 cm) wide on the heel side for forward and backward directions (this plate was removed before the trial). Participants’ feet were together for all left and right trials.

Precise timing of the release was obtained using inertial measurement units (IMU, Opal v2, APDM, Inc.) affixed to the participants’ lumbar spine, sternum, and feet and the administrator’s hand. For the purpose of this study, only the IMU data from the administrator’s hand was used. The release point (i.e., perturbation onset) was identified when the IMU on the administrator’s hand exceeded a movement threshold, defined as an acceleration magnitude >1.05 times gravity, indicating the release of the participant.^22^ All IMU data was sampled at 128 Hz.

### 1.3. EEG data analysis

EEG data were recorded from participants during the instrumented Push and Release using a 32-channel active electrode mobile Brain Vision LiveAmp recorder system (sampling frequency 250 Hz; impedance <25 kΩ) with a 24-bit A/D converter and online 20 kHz anti-aliasing low-pass filter and online reference of FCz. EEG data were synchronized with the IMU data using a 1-bit input TTL-pulse input trigger, recorded as an event marker in the EEG data. Data were processed offline using EEGLAB (Delorme & Makeig, 2004) with MATLAB. The raw EEG data were preprocessed in EEGlab using the BeMoBIL Pipeline.^25^ As part of the BeMoBIL preprocessing pipeline, line noise was removed using the Zapline plus plugin. Bad channels were detected and removed using the cleanrawdata plugin, interpolated and then full-rank average referenced. Preprocessed data were then high-pass filtered with a cut-off of 1.75 Hz and epoched -2 to 2 seconds around each perturbation (defined as the release time from the IMU data), given the discrete nature of “release” involved in the balance task.^26^ Adaptive mixture component analysis algorithm (AMICA) was used to identify maximally independent components (ICs).^27^ ICs that were identified by the algorithm as non-brain sources were further verified through visual inspection before removal. AMICA and evoked response analyses require different filtering, thus we then reverted to the BeMoBIL preprocessed unfiltered data to apply different filters before using the identified components to remove non-brain sources.

### Pre-perturbation cortical beta activity

To investigate anticipatory cortical activity and differences in pre-perturbation brain state between groups, ongoing pre-perturbation EEG activity was analyzed at the channel level (Cz) to capture midline -central preparatory activity. Pre-perturbation beta activity was extracted from the Cz electrode because anticipatory beta oscillations during standing balance are not source-localized to a dominant brain source but instead reflect broad sensorimotor network activity best captured at the central midline overlying the vertex. Beta activity (13-30Hz) is transient and burst-like,^13^ thus we analyzed the characteristics of individual beta burst-like events on a trial-by-trial basis. Spectral events that exceeded six times the median power of the whole time-frequency power matrix were identified as local maxima in the trial-by-trial time-frequency matrix using the SpectralEvents Toolbox by Shin et al.^13^ We extracted the beta event power and beta event duration of the most recent beta event and during the 1s time period prior to the onset of the balance release perturbation for each single trial. We computed the beta event amplitude and duration as the mean of the most recent event relative to the perturbation onset for all trials in each condition for each participant.

### Post-perturbation cortical N1 identification

The independent source component most likely to represent the N1 potential was identified by inspection of the event-related potential (ERP) waveform associated with each brain IC.^19^ For each participant, the single IC exhibiting the largest amplitude potential in the 50-250ms time period following the onset of the release was selected as the source-resolved N1 component. To validate the cortical origin and anatomical localization, equivalent current dipoles were fitted to the scalp projections of selected ICs using the DIPFIT plugin in EEGLAB (Oostenveld & Oostendorp, 2002) with a four-shell spherical head model and standard electrode positions.^19,28^ The N1 IC orientation was standardized to a common negative polarity to ensure consistency across participants. The source-resolved IC time series was low-pass filtered at 20 Hz and baseline subtracted (−150 to −50 ms).^19,26,29,30^ To ensure that the N1 peak was consistently identified within a participant across each EO and EC conditions, we calculated a grand-mean ERP by averaging the source-resolved IC time series across all perturbation trials and conditions. The N1 peak for the averaged ERP of each the EO and EC conditions was identified as the local minimum closest in time to the grand-mean N1 latency across both conditions. We extracted N1 peak latency and amplitude relative to perturbation onset (release time).

### 1.4. Statistical analyses Statistical Analysis

All statistical analyses were performed in R (version 4.3.3) using the lme4 and lmerTest packages.^31,32^ Given the small sample size and the preliminary nature of this study, we employed linear mixed-effects models (LMMs) to appropriately account for within-subject dependence arising from repeated measures per participant through the inclusion of a random intercept for participant. Given the hypothesis generating nature of the work, we present p-values continuously as a measure of compatibility with the null hypothesis (i.e., lower p-values are less compatible), descriptively present all data in our figures when applicable, and provide 95% confidence interval to show possible effect sizes in the population that are compatible with the data observed in the sample.^33^ Effect sizes were estimated as Hedge’s g to reduce upward bias in effect size estimates with small degrees of freedom.^34^ Effect sizes are reported with 95% confidence intervals derived from the Kenward-Roger degrees of freedom approximation. Note that even when p < 0.05 and the raw confidence interval excludes zero, the Hedges’ g confidence interval may include zero, reflecting the substantial additional uncertainty introduced when standardizing the effect in a small sample.

First, we explored group differences in pre-perturbation beta power and cortical N1 responses during balance recovery. Separate LMMs were used for each dependent variable of beta burst event metrics and N1 response amplitude and latency. Each model included group (mTBI, control), visual condition (eyes open, eyes closed), and their interaction as fixed effects, with a random intercept for participant. The group main effect tested whether mTBI participants exhibited differences in cortical activity metrics averaged across visual conditions. The visual condition main effect and the group-by-condition interaction were included to determine whether group differences were consistent across eyes-open and eyes-closed conditions. Estimated marginal means were computed for each group and contrasted to derive effect sizes.

We also explored whether mTBI had an effect on relationships between preparatory beta activity and evoked N1 responses during balance recovery. Separate LMMs were fit to test whether group moderated the association between the most recent beta event power and duration in predicting the N1 response outcomes of amplitude and duration. We explored the beta predictor-by-group interaction to see how much the strength or direction of the beta-N1 relationship differed between groups. Within-group Pearson correlations were also computed to characterize the direction of associations in each group. Given the exploratory nature and small sample, we did not apply statistical correction for the p-values presented, but all analyses are presented.^35^

## RESULTS

One control participant had a technical issue with recording the time to release event trigger and was excluded from all analyses. Groups were similar different in age and sex distribution.

Participants after mTBI reported higher symptom burden on the Post-Concussion Symptom Scale compared with controls and were assessed at a mean of 27 ± 13 days after injury (**Table 1**). People after mTBI tended to take longer to initiate a reactive step, but notably the time to stability in each group was quite similar.

### Participants after mTBI display lower cortical beta activity when preparing to recover balance

The mTBI group displayed lower mean preparatory beta burst power of all beta events compared to controls (Control: M = 6.92; mTBI: M = 4.37; F_1,7_ = 8.20, p = 0.024, mean difference=2.55, 95% CI [0.44, 4.65]; g = 1.23, 95% CI [−0.05, 2.51]), with a similar pattern for the most recent pre-perturbation burst power (Control: M = 7.26; mTBI: M = 4.39; F_1,7_ = 6.01, p = 0.044, mean difference = 2.86, 95% CI [0.10, 5.63]; g = 1.12, 95% CI [−0.17, 2.42]). Beta burst durations also tended to be shorter in the mTBI group, but this difference was more consistent with the null hypothesis (F_1,7_ = 2.76, p = 0.141, mean difference = 0.033, 95% CI [−0.014, 0.080]; g = 0.76, 95% CI [−0.42, 1.94]; see **Figure 1**). We did not see any evidence of statistically surprising differences between visual conditions or group-by-condition interactions for any beta measure (all p’s > 0.25).

**Figure 1.**
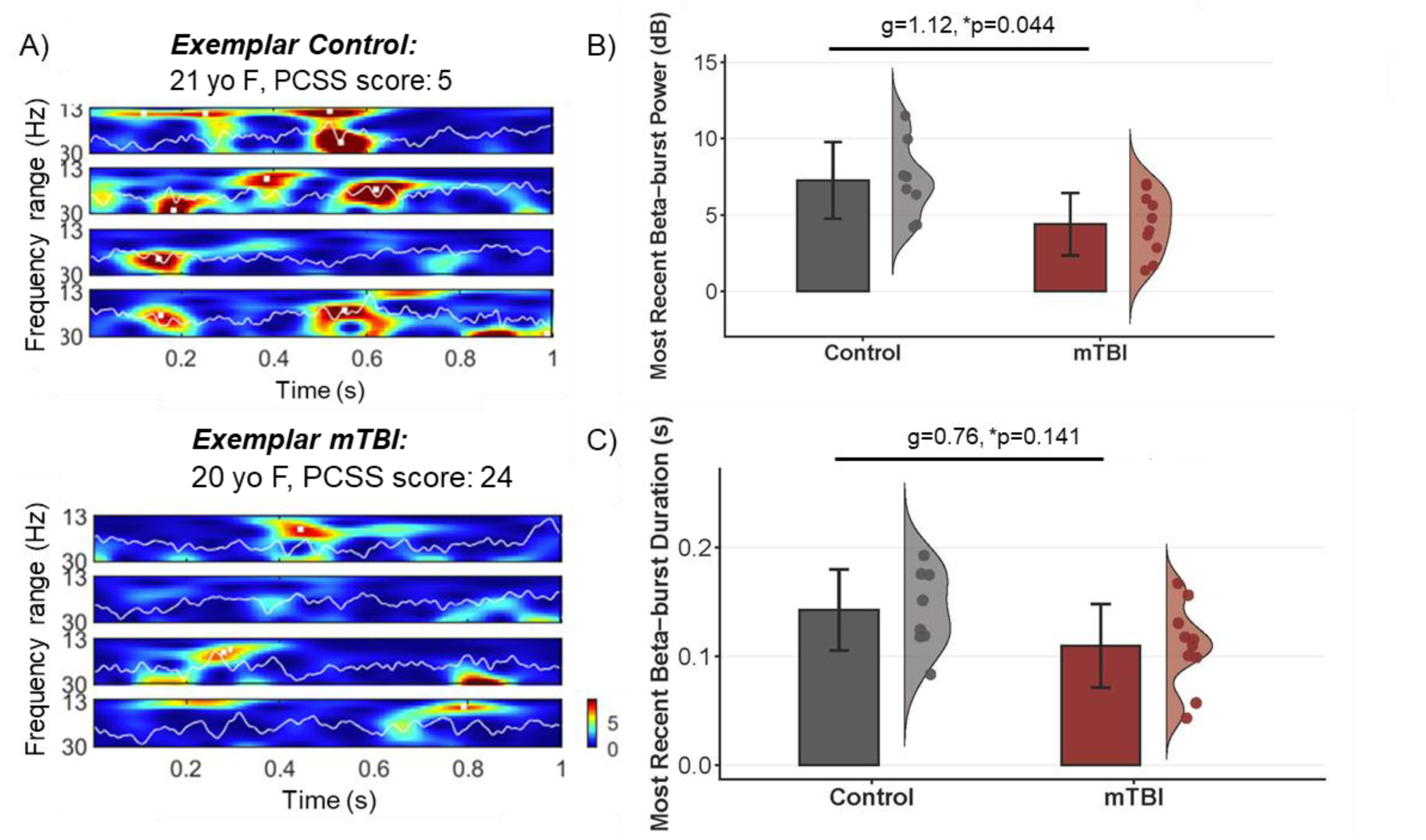
Preparatory cortical beta activity in participants after mTBI and controls. A) Beta event characteristics of 4 single trials in the 1s prior to the onset of the perturbation release. An individual with mTBI (bottom) had lower beta event power, and shorter beta event duration compared with the individual control (top). B) Group comparison of most recent pre-perturbation beta-burst power, collapsed across visual conditions. The mTBI group displayed lower beta-burst power compared with controls (p = 0.044, g = 1.12). C) Group comparison of most recent pre-perturbation beta-burst duration, collapsed across visual conditions. The mTBI group showed shorter beta-burst durations in the sample, but this difference was less surprising under the null hypothesis (p = 0.141, g = 0.76). Bars represent group mean ± standard deviation with individual participant data points overlaid.

### Cortical N1 responses occurred earlier during balance recovery after mTBI

The mTBI group exhibited shorter N1 latencies compared to controls (Control: M = 163.8 ms; mTBI: M = 101.2 ms; F_1, 7_ = 5.94, p = 0.045, mean difference = 62.6 ms, 95% CI [1.86, 123.37]; g = 3.11, 95% CI [−0.49, 6.72]), indicating earlier cortical evoked responses following perturbation onset. The evoked cortical N1 amplitude was numerically smaller in the mTBI group in the sample (Control: M = 7.82 µV; mTBI: M = 4.40 µV), but this difference was more consistent with the null hypothesis (F_1, 7_ = 2.75, p = 0.141, mean difference = 3.42 µV, 95% CI [−1.46, 8.30]; g = 2.39, 95% CI [−1.27, 6.11] (**Figure 2**). We did not see evidence of statistically surprising differences between visual conditions or group-by-condition interactions (all p’s > 0.37).

**Figure 2.**
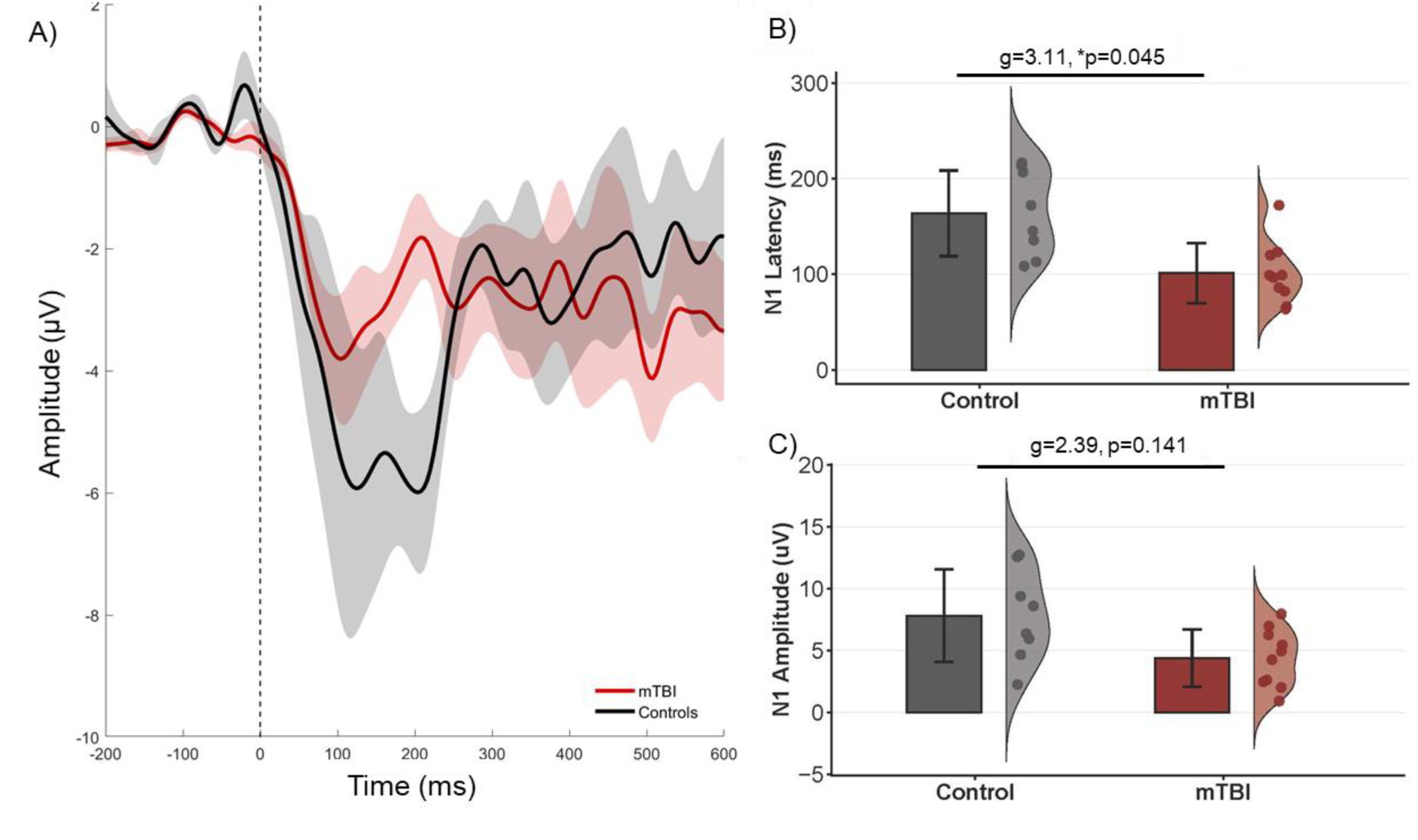
Evoked cortical N1activity in participants after mTBI and controls. A) Group mean N1 waveforms collapsed across conditions. Shaded region represents standard error of the mean (SEM) for each participant group. B) Group comparison of N1 latency, collapsed across conditions. The mTBI group exhibited earlier N1 latencies compared with controls (p = 0.045, g = 3.11). C) Group comparison of N1 amplitude, collapsed across visual conditions. The mTBI group showed numerically smaller N1 amplitudes in the sample, but this difference was less surprising under the null hypothesis (p = 0.141, g = 2.39. Bars represent group mean ± standard deviation with individual participant data points overlaid.

### Altered relationships between preparatory and evoked cortical activity during balance recovery after mTBI

Our preliminary data suggest that group may moderate the relationship between most recent beta-burst duration and N1 latency (interaction: F_1, 8.52_= 5.95, p = 0.039), such that longer burst durations were associated with shorter N1 latencies in the mTBI group (r = −0.77 95% CI [−0.94, −0.27], p = 0.010) but tended toward longer N1 latencies in controls (r = 0.69, 95% CI [−0.04, 0.94], p = 0.060). The same pattern was directionally consistent for most recent burst power predicting N1 latency, such that greater beta-burst power was associated with shorter N1 latencies in the mTBI group (r = −0.85, 95% CI [−0.96, −0.48], p = 0.002) and but not in controls (r = 0.34, 95% CI [−0.48, 0.84], p = 0.404), but there was considerable variability in the estimates, with no compelling evidence of a moderating effect on burst power (F_1, 10.06_ = 1.82, p = 0.207). Similarly, the relationship between beta-burst power and N1 Amplitude was fairly consistent between groups (F_1, 8.54_ = 1.46, p = 0.259), as was the relationship between beta-burst duration and N1 amplitude (F_1, 7.74_ = 2.46, p = 0.157).

There were no significant main effects of beta-burst power (F(1, 8.54) = 0.09, p = 0.770) or duration (F(1, 7.74) = 0.64, p = 0.448) on N1 amplitude when collapsed across groups. Additionally, although estimates of this relationship tended to be negative in both groups (**Figure 3**) there was considerable variance in these estimates as well: mTBI group (r = −0.53, 95% CI [−0.87, 0.15] for burst power; r = −0.37, 95% CI [−0.81, 0.34] for burst duration) and controls (r = −0.13, 95% CI [−0.77, 0.63] for burst power; r = −0.30, 95% CI [−0.83, 0.52] for burst duration).

**Figure 3.**
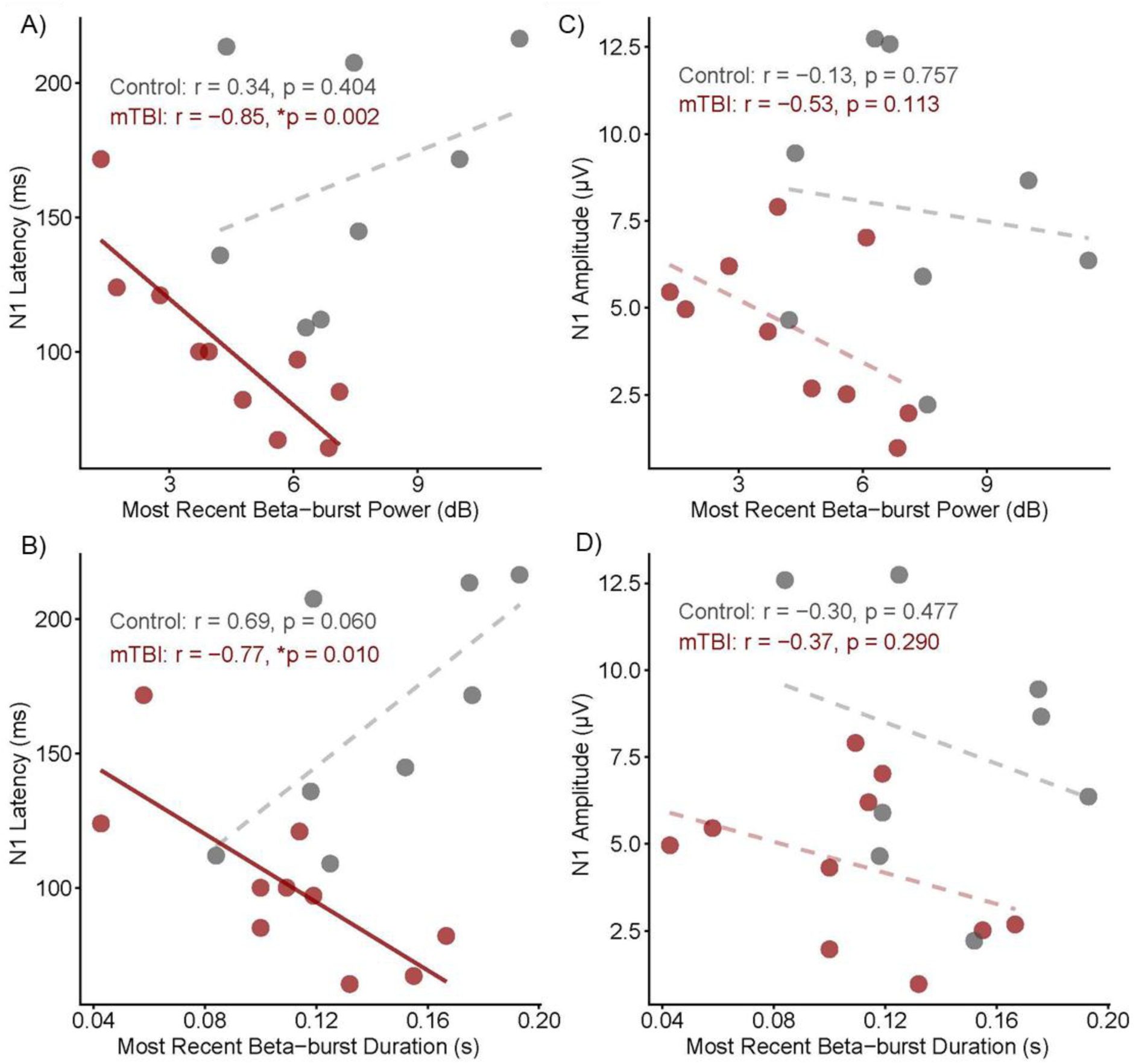
Relationships between preparatory and evoked cortical activity during balance recovery after mTBI and in controls. Each data point represents a visual condition for each participant, with separate regression lines for each group. A) Most recent pre-perturbation beta-burst power versus evoked cortical N1 response latency. Greater beta-burst power was associated with shorter N1 latencies in the mTBI group (r = −0.85, p = 0.002) with no clear relationship in controls (r=0.34). B) Longer beta-burst durations were associated with shorter evoked N1 response latencies in the mTBI group (r = −0.77, p = 0.010) but tended toward longer N1 latencies in controls (r = 0.69, p = 0.060). C) Most recent pre-perturbation beta-burst power versus N1 amplitude and D) Most recent pre-perturbation beta-burst duration versus N1 amplitude showed no relationships for either burst power or burst duration.

## DISCUSSION

The present study provides preliminary evidence that reactive balance control after mTBI is characterized by altered cortical activity during both the preparation for and execution of balance recovery. People in the sub-acute stage of mTBI recovery exhibited lower preparatory beta-burst power, earlier cortical N1 latencies, and altered coupling between preparatory and evoked cortical responses compared to age-matched controls. These findings suggest that mTBI may disrupt cortical inhibitory processes involved in some gating and response modulation prior to balance perturbations, as well as the temporal dynamics of cortical error detection during balance recovery. These preliminary neural signatures are consistent with clinical features commonly reported after mTBI, including sensory hypersensitivity,^15^ impaired executive function,^36–38^ and persistent balance dysfunction^7–9^ and may serve to guide future research directions in mTBI patient populations.

### Lower pre-perturbation beta-burst power in people after mTBI may reflect impaired sensory gating of somatosensory information during preparation for balance recovery

As shown in Figure 1, people after mTBI exhibited lower preparatory beta-burst power compared to controls regardless of visual condition. Beta oscillations reflect inhibitory cortical network activity and have been shown to suppress perception of somatosensory stimuli prior to movement.^13,28^ In neurologically-intact adults, this sensory gating process may prevent the central nervous system from being overwhelmed by predictable or irrelevant afferent feedback during movement preparation, allowing selective attention to task-relevant sensory information.^13^ Lower beta activity in people after mTBI may reflect a reduced capacity to filter predictable or task-irrelevant sensory information during balance preparation, which could interfere with the precision of subsequent motor responses. This interpretation is consistent with evidence of altered inhibitory network function after mTBI, including reduced cortical GABA concentrations in frontal regions during sub-acute recovery^39^ and in people with a history of repetitive head trauma.^40^ Clinically, sensory hypersensitivity is a symptom after TBI that persists in some individuals,^15^ and impaired sensory gating measured behaviorally has been implicated in the development of post-concussive symptoms.^41^ To our knowledge, the present findings provide the first preliminary evidence of impaired inhibitory cortical activity during whole-body sensorimotor preparation for balance recovery after brain injury, finding generally large effects but with considerable uncertainty: burst power g = 1.12, 95% CI = [−0.17, 2.42]; burst duration g = 0.76, 95% CI = [−0.42, 1.94]]. Future work with larger samples may examine relationships between beta-mediated sensory gating deficits and sensory sensitivity symptoms, for instance, for a two-sample test, assuming equal numbers of people with mTBI and controls, α=0.05, and that the more modest g=0.80 is a better reflection of the true effect size - that would require 68 people (34 per group) to achieve 90% statistical power.^42^ This sample size could be reduced through other power saving methods, e.g., collecting more events per participants to get a more reliable estimate of beta power/duration, but provides a low-end estimate of the sample size required to reliably test hypotheses about group differences in event-related beta in these populations. (Given that we observed a such a large effect in such a small sample, it is likely that g=0.80 is still an overestimate of the true effect size – see “the winner’s curse”).^43^

### Lower pre-perturbation beta-burst activity in people after mTBI may also reflect impaired response inhibition, a cortical process critical for guiding the precision and online adjustment of reactive balance responses

Effective reactive balance recovery requires both perceptual inhibition to rapidly attend to the most salient stimuli during a postural challenge, and motor inhibition to suppress or adjust pre-potent responses based on the environmental context.^44,45^ Beta-bursts in frontal and central cortical regions are associated with successful movement cancellation, with increased beta activity preceding suppression of unwanted motor responses.^46^ Bolton et al. (2026) recently extended this response inhibition framework to reactive balance, demonstrating that higher pre-perturbation beta power was associated with successful suppression of a prepotent stepping response during a modified lean-and-release paradigm similar to the present study in neurotypical adults.^47^ Response inhibition as a component of cognitive executive function has been identified as a prevalent cognitive impairment after mTBI,^36–38^ yet its role in reactive balance control after brain injury has not been examined. These preliminary findings suggest that, in addition to sensory gating, the lower preparatory beta-burst activity may also reflect diminished cortical inhibitory network readiness that could compromise the capacity to filter competing sensory information and adjust stepping responses with precision during balance recovery after mTBI.

### Earlier cortical N1 latencies in people after mTBI may reflect altered temporal dynamics of balance error signaling that distinguish mTBI from other neurologic conditions

In people after stroke, delayed cortical N1 responses are associated with delayed step initiation and lower clinical balance function, implicating slowed cortical sensorimotor error detection and processing as a neural substrate underpinning reactive balance deficits.^29,48^ In contrast, people after mTBI in the present study exhibited significantly earlier N1 latencies (∼101 ms vs ∼164 ms in controls), a pattern inconsistent with slowed processing speed and instead suggesting altered temporal dynamics of cortical error detection. One interpretation is that earlier N1 latencies reflect greater balance challenge, as cortical responses can occur earlier under more demanding balance conditions.^49–51^ The mTBI group may have experienced the push-and-release paradigm as more destabilizing, eliciting earlier cortical error signals. However, earlier cortical error detection did not translate to faster motor execution in this cohort; people after mTBI trended toward longer, not shorter, step latencies. This dissociation suggests that the coupling between cortical error signals and downstream motor responses may be disrupted after mTBI. Additionally, N1 amplitudes tended to be smaller in mTBI, which is notable given that larger cortical responses are typically associated with worse balance performance in neurotypical adults.^49,51^ Future studies are needed to test whether earlier and smaller cortical N1 responses in people after mTBI reflect a dampened capacity for cortical error detection that may limit balance learning and adaptation.

### Altered relationships between preparatory and evoked cortical activity during balance recovery after mTBI are consistent with a shift in neural strategy involving preparation and execution of balance recovery

As shown in Figure 3, the relationship between preparatory beta burst duration and N1 latency was different between people after mTBI and controls. This preliminary finding suggests that the typical coupling between preparatory inhibitory processes and subsequent cortical error detection may be disrupted after mTBI. Recent work from our group demonstrated that reactive balance deficits differ based on symptom chronicity after mTBI.^7^ People with acute symptoms exhibit delayed step initiation but normal time to stability, while those with subacute and chronic symptoms show normal step latencies but took a longer time to regain stability.^7^ The differences in reactive balance recovery strategy as a function of mTBI chronicity may suggest a shift from slower, more cautious responses toward faster but less precise stepping strategies over the course of recovery. The present cohort was in the sub-acute stage of recovery (∼27 days post-injury), a period during which this transition may emerge. In neurotypical adults, preparatory inhibitory activity may appropriately coordinate with subsequent error detection processes, facilitating effective transitions from postural maintenance to balance recovery.

Disruption of this coupling after mTBI could contribute to the dissociation between rapid step initiation and impaired balance stabilization observed in people with persisting symptoms. These preliminary findings suggest that quantifying both the strength and coordination of cortical preparatory and reactive processes may provide insight into neural mechanisms underlying balance dysfunction and could inform rehabilitation strategies targeting preparatory and execution components of reactive balance control after mTBI. For instance, the relationship between N1 latency and beta burst power/duration were generally negative in people with mTBI (r’s =-0.85 to - 0.77) and positive in controls (r’s = 0.34 to 0.69). If we assume more modest differences (i.e., true mTBI r=-0.6 and true control r=0.30), then it would require N=48, 24 per group, to reliably detect the difference between these two independent correlations, assuming α=0.05, with 90% statistical power.

## CONCLUSION

The present preliminary findings show altered cortical activity during distinct preparatory and execution phases of balance recovery and highlight the importance of neuroimaging modalities with high temporal resolution, such as EEG, for assessing neural signatures of reactive balance dynamics. The results in the present small cohort are intended as preliminary, hypothesis-generating observations to guide future research directions. Larger mechanistic studies are needed to determine whether cortical signatures of preparatory and execution elements of reactive balance control predict clinical outcomes, track symptom recovery, or show modulation with rehabilitation interventions. Identifying neurophysiological markers relevant to balance preparation and execution may ultimately support precision-based approaches to assessment and treatment for people after mTBI.

## Data Availability

Data produced in the present study are available upon reasonable request to the authors.

## ACKNOWLEDGEMENTS

The authors acknowledge Breanna Dumke, Nicholas Kreter, Cameron Stukel, and Claire Rogers for their assistance with data collection and Jacquelyn Sertic and Kathleen Gurin for their assistance with initial steps of EEG analysis.

## Author contributions

JP and PF conceived of the present research question and designed this study analysis plan. PF conceived of the experimental design. PF and KL supervised the data collection. PF and KL obtained the primary funding to support this study. JP performed the EEG data analyses, constructed the figures, and drafted the first version of this manuscript. All authors discussed the results and contributed to the final manuscript.

## REFERENCES

1. Nelson LD, Temkin NR, Dikmen S, et al. Recovery After Mild Traumatic Brain Injury in Patients Presenting to US Level I Trauma Centers: A Transforming Research and Clinical Knowledge in Traumatic Brain Injury (TRACK-TBI) Study. JAMA Neurol. 2019;76(9):1049–1059. doi:10.1001/jamaneurol.2019.1313

2. Guskiewicz KM. Balance assessment in the management of sport-related concussion. Clin Sports Med. 2011;30(1):89–102, ix. doi:10.1016/j.csm.2010.09.004

3. Quatman-Yates CC, Hunter-Giordano A, Shimamura KK, et al. Physical Therapy Evaluation and Treatment After Concussion/Mild Traumatic Brain Injury. Journal of Orthopaedic & Sports Physical Therapy. 2020;50(4):CPG1-CPG73. doi:10.2519/jospt.2020.0301

4. Irimia A, Wang B, Aylward SR, et al. Neuroimaging of structural pathology and connectomics in traumatic brain injury: Toward personalized outcome prediction. NeuroImage: Clinical. 2012;1(1):1–17. doi:10.1016/j.nicl.2012.08.002

5. Povlishock JT, Christman CW. The pathobiology of traumatically induced axonal injury in animals and humans: a review of current thoughts. J Neurotrauma. 1995;12(4):555–564. doi:10.1089/neu.1995.12.555

6. Ferrazzano PA, Rebsamen S, Field AS, et al. MRI and Clinical Variables for Prediction of Outcomes After Pediatric Severe Traumatic Brain Injury. JAMA Netw Open. 2024;7(8):e2425765. doi:10.1001/jamanetworkopen.2024.25765

7. Monoli C, Johnson PK, Morris AJ, Pelo RM, Dibble LE, Fino PC. Reactive Balance Deficits Differ in People After Mild Traumatic Brain Injury Based on Chronicity of Self-Reported Symptoms. Neurorehabil Neural Repair. Published online January 26, 2026:15459683251412303. doi:10.1177/15459683251412303

8. Monoli C, Morris AJ, Crofts R, et al. Acute and Longitudinal Effects of Concussion on Reactive Balance in Collegiate Athletes. Neurorehabil Neural Repair. 2025;39(4):263–273. doi:10.1177/15459683241309569

9. Pan T, Liao K, Roenigk K, Daly JJ, Walker MF. Static and dynamic postural stability in veterans with combat-related mild traumatic brain injury. Gait Posture. 2015;42(4):550–557. doi:10.1016/j.gaitpost.2015.08.012

10. Horak FB, Jacobs JV. Cortical control of postural responses. Journal of Neural Transmission. 2007;114(10):1339–1348. doi:10.1007/s00702-007-0657-0.Cortical

11. Maki BE, McIlroy WE. Cognitive demands and cortical control of human balance-recovery reactions. Journal of Neural Transmission. 2007;114(10):1279–1296. doi:10.1007/s00702-007-0764-y

12. Martini DN, Mancini M, Antonellis P, et al. Prefrontal Cortex Activity During Gait in People With Persistent Symptoms After Concussion. Neurorehabil Neural Repair. 2024;38(5):364–372. doi:10.1177/15459683241240423

13. Shin H, Law R, Tsutsui S, Moore CI, Jones SR. The rate of transient beta frequency events predicts behavior across tasks and species. Kajikawa Y, ed. eLife. 2017;6:e29086. doi:10.7554/eLife.29086

14. Mirdamadi JL, Ting LH, Borich MR. Distinct Cortical Correlates of Perception and Motor Function in Balance Control. J Neurosci. 2024;44(15):e1520232024. doi:10.1523/JNEUROSCI.1520-23.2024

15. Callahan ML, Lim MM. Sensory Sensitivity in TBI: Implications for Chronic Disability. Curr Neurol Neurosci Rep. 2018;18(9):56. doi:10.1007/s11910-018-0867-x

16. Payne AM, Schmidt NB, Meyer A, Hajcak G. The balance N1 is larger in anxious children and associated with the error-related negativity. Biological Psychiatry: Global Open Science. 2024;0(0). doi:10.1016/j.bpsgos.2024.100393

17. Marlin A, Mochizuki G, Staines WR, McIlroy WE. Localizing evoked cortical activity associated with balance reactions: does the anterior cingulate play a role? Journal of Neurophysiology. 2014;111(12):2634–2643. doi:10.1152/jn.00511.2013

18. Mierau A, Hülsdünker T, Strüder HK. Changes in cortical activity associated with adaptive behavior during repeated balance perturbation of unpredictable timing. Front Behav Neurosci. 2015;9. doi:10.3389/fnbeh.2015.00272

19. Solis-Escalante T, Stokkermans M, Cohen MX, Weerdesteyn V. Cortical responses to whole-body balance perturbations index perturbation magnitude and predict reactive stepping behavior. Eur J Neurosci. Published online September 15, 2020. doi:10.1111/ejn.14972

20. Payne AM, Ting LH, Hajcak G. Do sensorimotor perturbations to standing balance elicit an error-related negativity? Psychophysiology. 2019;56(7):1–12. doi:10.1111/psyp.13359

21. Franchignoni F, Horak F, Godi M, Nardone A, Giordano A. Using psychometric techniques to improve the Balance Evaluation Systems Test: the mini-BESTest. J Rehabil Med. 2010;42(4):323–331. doi:10.2340/16501977-0537

22. Morris A, Fino NF, Pelo R, et al. Interadministrator Reliability of a Modified Instrumented Push and Release Test of Reactive Balance. J Sport Rehabil. 2022;31(4):517–523. doi:10.1123/jsr.2021-0229

23. Petersell TL, Quammen DL, Crofts R, et al. Instrumented Static and Reactive Balance in Collegiate Athletes: Normative Values and Minimal Detectable Change. J Athl Train. 2024;59(6):608–616. doi:10.4085/1062-6050-0403.23

24. Kreter N, Rogers CL, Fino PC. Anticipatory and reactive responses to underfoot perturbations during gait in healthy adults and individuals with a recent mild traumatic brain injury. Clinical Biomechanics. 2021;90:105496. doi:10.1016/j.clinbiomech.2021.105496

25. Klug M, Jeung S, Wunderlich A, et al. The BeMoBIL Pipeline for automated analyses of multimodal mobile brain and body imaging data. bioRxiv. Preprint posted online October 6, 2022:2022.09.29.510051. doi:10.1101/2022.09.29.510051

26. Mirdamadi JL, Poorman A, Munter G, et al. Excellent test-retest reliability of perturbation-evoked cortical responses supports feasibility of the balance N1 as a clinical biomarker. Journal of Neurophysiology. 2025;133(3):987–1001. doi:10.1152/jn.00583.2024

27. Delorme A, Palmer J, Onton J, Oostenveld R, Makeig S. Independent EEG Sources Are Dipolar. PLOS ONE. 2012;7(2):e30135. doi:10.1371/journal.pone.0030135

28. Mirdamadi JL, Ting LH, Borich MR. Distinct Cortical Correlates of Perception and Motor Function in Balance Control. J Neurosci. 2024;44(15):e1520232024. doi:10.1523/JNEUROSCI.1520-23.2024

29. Palmer JA, Payne AM, Mirdamadi JL, Ting LH, Borich MR. Delayed Cortical Responses During Reactive Balance After Stroke Associated With Slower Kinetics and Clinical Balance Dysfunction. Neurorehabil Neural Repair. Published online September 27, 2024:15459683241282786. doi:10.1177/15459683241282786

30. Payne AM, McKay JL, Ting LH. The cortical N1 response to balance perturbation is associated with balance and cognitive function in different ways between older adults with and without Parkinson’s disease. Cereb Cortex Commun. 2022;3(3):tgac030. doi:10.1093/texcom/tgac030

31. Bates D, Mächler M, Bolker B, Walker S. Fitting Linear Mixed-Effects Models Using lme4. Journal of Statistical Software. 2015;67:1–48. doi:10.18637/jss.v067.i01

32. Kuznetsova A, Brockhoff PB, Christensen RHB. lmerTest Package: Tests in Linear Mixed Effects Models. Journal of Statistical Software. 2017;82:1–26. doi:10.18637/jss.v082.i13

33. Greenland S, Senn SJ, Rothman KJ, et al. Statistical tests, P values, confidence intervals, and power: a guide to misinterpretations. Eur J Epidemiol. 2016;31:337–350. doi:10.1007/s10654-016-0149-3

34. Hedges LV. Distribution Theory for Glass’s Estimator of Effect size and Related Estimators. Journal of Educational Statistics. 1981;6(2):107–128. doi:10.3102/10769986006002107

35. Bender R, Lange S. Adjusting for multiple testing--when and how? J Clin Epidemiol. 2001;54(4):343–349. doi:10.1016/s0895-4356(00)00314-0

36. Xu B, Sandrini M, Levy S, et al. Lasting deficit in inhibitory control with mild traumatic brain injury. Sci Rep. 2017;7(1):14902. doi:10.1038/s41598-017-14867-y

37. DeHaan A, Halterman C, Langan J, et al. Cancelling planned actions following mild traumatic brain injury. Neuropsychologia. 2007;45(2):406–411. doi:10.1016/j.neuropsychologia.2006.06.008

38. Shen IH, Lin YJ, Chen CL, Liao CC. Neural Correlates of Response Inhibition and Error Processing in Individuals with Mild Traumatic Brain Injury: An Event-Related Potential Study. J Neurotrauma. 2020;37(1):115–124. doi:10.1089/neu.2018.6122

39. Yasen AL, Smith J, Christie AD. Glutamate and GABA concentrations following mild traumatic brain injury: a pilot study. J Neurophysiol. 2018;120(3):1318–1322. doi:10.1152/jn.00896.2017

40. Kim GH, Kang I, Jeong H, et al. Low Prefrontal GABA Levels Are Associated With Poor Cognitive Functions in Professional Boxers. Front Hum Neurosci. 2019;13:193. doi:10.3389/fnhum.2019.00193

41. Kumar S, Rao SL, Nair RG, Pillai S, Chandramouli BA, Subbakrishna DK. Sensory gating impairment in development of post-concussive symptoms in mild head injury. Psychiatry Clin Neurosci. 2005;59(4):466–472. doi:10.1111/j.1440-1819.2005.01400.x

42. Faul F, Erdfelder E, Buchner A, Lang AG. Statistical power analyses using G*Power 3.1: Tests for correlation and regression analyses. Behavior Research Methods. 2009;41(4):1149–1160. doi:10.3758/BRM.41.4.1149

43. Button KS, Ioannidis JPA, Mokrysz C, et al. Power failure: why small sample size undermines the reliability of neuroscience. Nat Rev Neurosci. 2013;14(5):365–376. doi:10.1038/nrn3475

44. Bolton DAE, Richardson JK. Inhibitory Control and Fall Prevention: Why Stopping Matters. Front Neurol. 2022;13. doi:10.3389/fneur.2022.853787

45. Bolton DAE, Mansour M. A Modified Lean and Release Technique to Emphasize Response Inhibition and Action Selection in Reactive Balance. J Vis Exp. 2020;(157). doi:10.3791/60688

46. Wessel JR. β-Bursts Reveal the Trial-to-Trial Dynamics of Movement Initiation and Cancellation. J Neurosci. 2020;40(2):411–423. doi:10.1523/JNEUROSCI.1887-19.2019

47. Bolton DAE, Beethe AZ, Harper SA, McNickle E, Whelan R, Ruddy KL. Establishing a Neural Marker for Inhibitory Control During Balance Recovery. Psychophysiology. 2026;63(1):e70208. doi:10.1111/psyp.70208

48. Tsai SY, Payne AM, Mirdamadi JL, Ting LH, Borich MR, Palmer JA. Slower reactive stepping kinematics are associated with lower clinical balance function and delayed cortical evoked responses under challenging balance conditions after stroke. medRxiv. Preprint posted online December 19, 2025:2025.12.18.25342582. doi:10.64898/2025.12.18.25342582

49. Ghosn NJ, Palmer JA, Borich MR, Ting LH, Payne AM. Cortical Beta Oscillatory Activity Evoked during Reactive Balance Recovery Scales with Perturbation Difficulty and Individual Balance Ability. Brain Sci. 2020;10(11). doi:10.3390/brainsci10110860

50. Parr JVV, Mills R, Kal E, Bronstein AM, Ellmers TJ. A “Conscious” Loss of Balance: Directing Attention to Movement Can Impair the Cortical Response to Postural Perturbations. J Neurosci. 2024;44(48). doi:10.1523/JNEUROSCI.0810-24.2024

51. Payne AM, Ting LH. Worse balance is associated with larger perturbation-evoked cortical responses in healthy young adults. Gait & Posture. 2020;80:324–330.

